# Human biodistribution and radiation dosimetry of the demyelination tracer [^18^F]3F4AP

**DOI:** 10.1101/2022.05.21.22275275

**Authors:** Pedro Brugarolas, Moses Q. Wilks, Jacqueline Noel, Julia-Ann Kaiser, Danielle R. Vesper, Karla M. Ramos-Torres, Nicolas J. Guehl, Marina T. Macdonald-Soccorso, Peter A. Rice, Daniel L. Yokell, Ruth Lim, Marc D. Normandin, Georges El Fakhri

## Abstract

**Purpose:** [^18^F]3F4AP is a novel PET radiotracer that targets voltage-gated potassium (K^+^) channels and has shown promise for imaging demyelinated lesions in animal models of neurological diseases. This study aimed to evaluate the biodistribution, safety and radiation dosimetry of [^18^F]3F4AP in healthy human volunteers.

**Methods:** Four healthy volunteers (2 female) underwent a 4-hour dynamic PET scan from cranial vertex to mid-thigh using multiple bed positions after administration of 368 ± 17.9 MBq (9.94 ± 0.48 mCi) of [^18^F]3F4AP was produced in a cGMP laboratory under IND authorization from the FDA. Volumes of interest for relevant organs were manually drawn guided by the CT and PET images and time-activity curves (TACs) were extracted. Radiation dosimetry was estimated from the integrated TACs using OLINDA software. Safety assessments included measuring vital signs immediately before and after the scan, monitoring for adverse events, as well as obtaining a comprehensive metabolic panel (CMP) and electrocardiogram (ECG) within 30 days before and after the scan. The study was approved by the institutional review board (IRB) and registered at clinicaltrials.gov (NCT04710550) before commencement.

**Results:** [^18^F]3F4AP distributed throughout the body with the highest levels of activity in the kidneys, urinary bladder, stomach, liver, spleen and brain and with low accumulation in muscle and fat. The tracer cleared quickly from circulation and from most organs. The clearance of the tracer was noticeably faster than previously reported clearance in nonhuman primates (NHPs). The average effective dose (ED) across all subjects was 12.1 ± 2.2 µSv/MBq, which is lower than the estimated ED from the NHP studies (21.6 ± 0.6 µSv/MBq) as well as the ED of other fluorine-18 radiotracers such as [^18^F]FDG (∼20 µSv/MBq). No differences in ED between males and females were observed. No substantial changes in safety assessments or adverse events were recorded.

**Conclusion:** The biodistribution and radiation dosimetry of [^18^F]3F4AP in humans is reported for the first time. The average total ED across four subjects was lower than most ^18^F-labeled PET tracers. The tracer and study procedures were well tolerated and no adverse events occurred.

## Background

[^18^F]3-fluoro-4-aminopyridine ([^18^F]3F4AP) is a radiofluorinated analog of the multiple sclerosis (MS) drug 4-aminopyridine (dalfampridine, 4AP)[1] (**Fig. 1**). Similar to 4AP, [^18^F]3F4AP binds to voltage-gated K^+^ channels (K_v_1 family) in demyelinated axons and has been proposed as a PET tracer for imaging demyelinated lesions in the brain[2-4]. In demyelinated lesions, axonal K^+^ channels K_v_1.1 and K_v_1.2, which are normally buried under the myelin sheath and confined to the juxtaparanodal regions of the axons, become exposed and increase in expression[2-4]. This aberrant expression of K^+^ channels results in excessive efflux of intracellular K^+^ ions and impaired axonal conduction, thus causing neurological deficits in MS and other demyelinating diseases. The FDA-approved drug 4AP binds to and blocks the K^+^ channels in demyelinated axons, reducing the abnormal efflux of K^+^ ions from axons and partially restoring conduction[5-9]. Given the increase in axonal K^+^ channel expression and the ability of 4AP to bind to these channels, [^18^F]3F4AP, a radiofluorinated analog of 4AP, was proposed as a PET tracer for demyelination[1, 10]. [^18^F]3F4AP is similar to 4AP in that it can enter the brain by passive diffusion and bind to K^+^ channels in demyelinated axons. Previous studies in rodents showed that [^18^F]3F4AP can be used to detect lesions in a rat model of demyelination using PET[1]. Additional studies in rhesus macaques showed that [^18^F]3F4AP has suitable properties for imaging primate brains including high brain penetration, fast kinetics, minimal plasma protein binding and high metabolic stability[11]. Furthermore, PET imaging of a monkey with a small focal traumatic brain injury sustained three years prior to imaging showed excellent sensitivity to the lesion[11]. These findings have prompted us to translate [^18^F]3F4AP to human research studies (Clinicaltrials.gov identifiers: NCT04699747, NCT04710550). As the first step in the evaluation of [^18^F]3F4AP in human subjects, we set out to assess the whole-body biodistribution, safety and radiation dosimetry in healthy human volunteers. We also compare these results with previous findings in nonhumans primates.

**Fig. 1.**
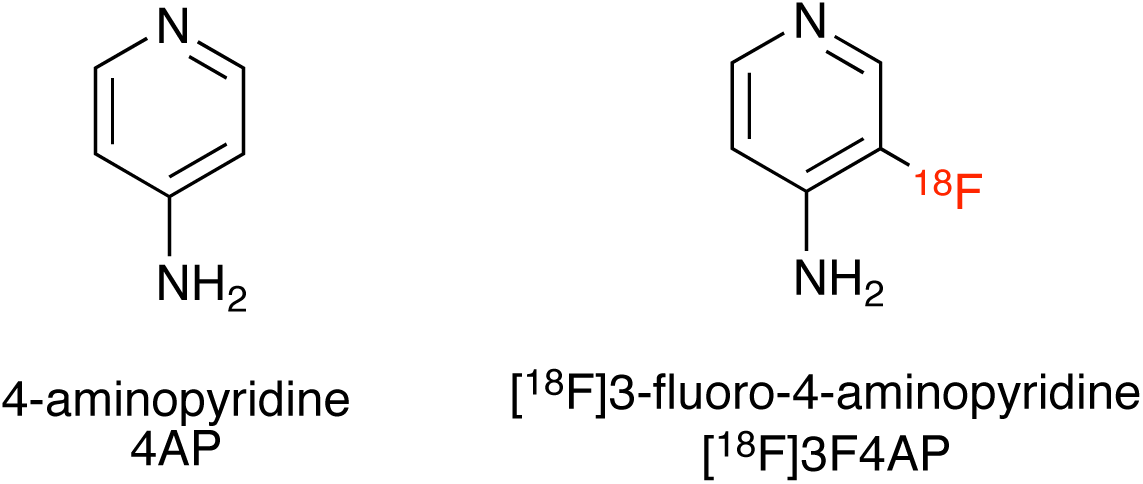
Chemical structures of 4-aminopyridine and [^18^F]3-fluoro-4-aminopyridine

## Methods

### Compliance

the study was approved by the Institutional Review Board at the Massachusetts General Hospital and registered in clinicaltrials.gov prior to initiation of the study (NCT04710550). [^18^F]3F4AP was administered under an investigational new drug (IND) authorization from the U.S. Food and Drug Administration (FDA).

### Subjects

four healthy volunteers (2 females, 2 males) participated in the study after providing informed consent. Participating criteria included adults between 18 and 65 years old with no history of brain disease or other serious condition, no contraindications to PET/CT such as severe claustrophobia, accumulated annual radiation dose less than 30 mSv, not taking dalfampridine containing drugs, normal results on a complete metabolic panel (CMP) blood test, normal results on electrocardiogram (ECG) and being able to provide informed consent. No subjects were excluded on the bases of sex, ethnicity or race.

### Safety assessments

vital signs including blood pressure, oxygen saturation and body temperature were collected immediately before and after completion of the scan. Subjects were queried about their level of comfort during and after the scan and were told to report any adverse events on the days following the scan. Within 30 days after completion of the scan, the subjects received a second CMP and a second ECG.

### Radiotracer production

[^18^F]3F4AP synthesis was performed using a Neptis ORA synthesizer as recently communicated[12]. The synthesis method is based on the previous report by Basuli et al[13]. The tracer was purified using a semiprep HPLC column (Waters XBridge C18 10 × 250 mm) using 10 mM sodium phosphate (pH 8) mobile phase containing 5% ethanol. The HPLC fraction containing the product (approx. 10-11 min) was diluted with 10 mL of 0.9% sodium chloride for injection, USP, and passed through a 0.22 µm sterilizing PES filter into a vented 30 mL sterile empty vial. The product vial was visually inspected and quality control was performed to FDA and USP standards for chemical identity and purity, radiochemical purity, pH, radionuclidic identity, residual solvents, sterility and bacterial endotoxins. The sterile filter used was tested for integrity. The dose was released for injection after passing all quality control tests except for sterility, which was performed after release. All of the batches of [^18^F]3F4AP used in this study met all product specifications, including sterility test.

### Radiotracer administration

the dose was drawn into a syringe, measured and administered intravenously as a 1 min infusion through a catheter in the hand or arm. After administration of the dose, the catheter was flushed with 10 mL of saline and the residual activity in the syringe and catheter measured to calculate the injected dose.

### Image acquisition protocol

imaging was performed on a GE Discovery MI PET/CT scanner. Subjects were positioned on the bed of the scanner and a low dose CT from cranial vertex to mid-thigh was acquired. Based on the CT images, seven bed positions with overlapping edges were selected for PET acquisition (25 cm per bed position with 2.5-5 cm overlap on each end). PET images were acquired over a period of four hours with two 15 min breaks at approximately 90 min and 160 min after tracer administration. Subjects were encouraged to void their bladder to eliminate radioactive urine during the breaks. After each break, a low dose CT was performed for anatomical reference and attenuation correction. The PET acquisition protocol consisted of a series of static images at each bed position of increasing duration starting upon administration of the tracer. The full acquisition protocol was as follows: low dose CT, 4 passes × 1 min PET acquisition per bed position, 4 passes × 2 min/bed, break, 2 passes × 4 min/bed, break, low dose CT, 1 pass × 7 min/bed position. After completion of the scan, the PET data was reconstructed using the scanner’s VUE Point HD reconstruction algorithm with 17 subsets and 3 iterations with corrections for scatter, attenuation, deadtime, random coincidence and scanner normalization.

### Blood sampling

3-5 mL venous blood samples were taken at approximately 8, 30, 60, 90, 135, 165 and 240 min post-injection. Blood samples’ radioactivity was counted using a calibrated gamma counter.

### Image analysis and dosimetry calculations

the CT and PET images were coregistered to the first CT images. Based on the CT and PET images, volumes of interest (VOIs) were manually drawn for the following tissues: adrenals, brain, breasts, gall bladder, small intestine, upper and lower large intestine, stomach, heart contents, heart muscle, kidney, liver, lung, muscle, ovaries, pancreas, red marrow, trabecular and cortical bone, spleen, testes, thymus, thyroid, urinary bladder and uterus. Time-activity curves (TACs) were extracted for each VOI and uncorrected for decay. TACs were extrapolated to ten half-lives after injection by assuming that any further decline in radioactivity occurred only due to physical decay with no biological clearance. Radiation dosimetry and effective dose were calculated from the integrated TACs using OLINDA software.

## Results

### Participant characteristics and radiotracer injection

The demographics of the volunteers are shown in **Table 1**. The study included two males and two females between the ages of 24 and 59 years old. Three out of the four subjects self-reported to be white and one reported more than one race. The body mass index of the participants ranged from 21.2 to 33. Each subject received an intravenous injection of [^18^F]3F4AP (target activity 370 ± 37 MBq) formulated in ∼10 mL of saline (**Table 2**). The molar activity of the tracer at the time of injection was greater than 370 GBq/µmol (1 Ci/µmol) as specified on the IND resulting in an injected mass of 3F4AP below 10 microgram, which is well below the pharmacologically active dose of 4AP (10 mg bid).

**Table 1.** Demographics of study participants

| Participant | Sex | Age range (years) | Race/ethnicity | Weight kg (lb) | Height cm (in) | BMI |
| --- | --- | --- | --- | --- | --- | --- |
| P1 | Female | 40-45 | Caucasian | 79.4 (175) | 155 (61) | 33 |
| P2 | Male | 40-45 | More than one race | 73.5 (162) | 175 (69) | 24 |
| P3 | Male | 55-60 | Caucasian | 61.2 (135) | 170 (67) | 21.2 |
| P4 | Female | 20-25 | Caucasian | 61.2 (135) | 163 (64) | 23 |

**Table 2.** Injected activity and effective dose for each participant

| Participant | Injected Activity MBq (mCi) | Effective Dose mSv |
| --- | --- | --- |
| P1 | 378.14 (10.22) | 4.20 |
| P2 | 381.84 (10.32) | 4.89 |
| P3 | 342.25 (9.25) | 3.29 |
| P4 | 380.73 (10.29) | 5.60 |

**Table 3.** Organ dosimetry calculations. Values are mean ± SD of 4 subjects.

| Target Organ | Residence time (h) | Organ Dose ($\mu\text{Gy}/\text{MBq}$) | Contribution to ED ($\mu\text{Sv}/\text{MBq}$) |
| --- | --- | --- | --- |
| Adrenals | $0.0007 \pm 0.0002$ | $14.6 \pm 3.6$ | $0.073 \pm 0.018$ |
| Brain | $0.031 \pm 0.004$ | $6.7 \pm 0.6$ | $0.034 \pm 0.003$ |
| Breasts | $0.004 \pm 0.002$ | $4.4 \pm 1.1$ | $0.221 \pm 0.055$ |
| Gallbladder | $0.0008 \pm 0.0004$ | $8.3 \pm 1.0$ | 0 |
| Lower large intestine | $0.0058 \pm 0.0007$ | $8.9 \pm 1.6$ | $1.064 \pm 0.196$ |
| Small intestine | $0.017 \pm 0.011$ | $8.3 \pm 2.9$ | $0.042 \pm 0.015$ |
| Stomach | $0.024 \pm 0.009$ | $14.6 \pm 3.6$ | $1.750 \pm 0.429$ |
| Upper large intestine | $0.0052 \pm 0.0017$ | $7.2 \pm 1.8$ | $0.036 \pm 0.009$ |
| Heart | $0.0106 \pm 0.0015$ | $54.6 \pm 4.0$ | 0 |
| Kidneys | $0.039 \pm 0.014$ | $28.6 \pm 9.3$ | $0.143 \pm 0.046$ |
| Liver | $0.106 \pm 0.018$ | $18.4 \pm 1.7$ | $0.922 \pm 0.086$ |
| Lungs | $0.015 \pm 0.003$ | $8.4 \pm 0.2$ | $1.003 \pm 0.027$ |
| Muscle | $0.52 \pm 0.30$ | $7.0 \pm 2.3$ | $0.035 \pm 0.012$ |
| Ovaries | $0.0007 \pm 0.0005$ | $14.9 \pm 4.5$ | $2.965 \pm 0.898$ |
| Pancreas | $0.0044 \pm 0.0006$ | $15.5 \pm 2.3$ | $0.077 \pm 0.011$ |
| Red Marrow | $0.052 \pm 0.020$ | $8.6 \pm 2.1$ | $1.034 \pm 0.248$ |
| Trabecular bone | $0.015 \pm 0.004$ | $13.2 \pm 1.6$ | $0.132 \pm 0.016$ |
| Cortical bone | $0.086 \pm 0.024$ | $2.6 \pm 0.6$ | $0.026 \pm 0.006$ |
| Spleen | $0.009 \pm 0.002$ | $15.0 \pm 3.4$ | $0.075 \pm 0.017$ |
| Testes | $0.0015 \pm 0.004$ | $9.8 \pm 2.2$ | $1.950 \pm 0.438$ |
| Thymus | $0.0006 \pm 0.0001$ | $11.7 \pm 1.6$ | $0.059 \pm 0.008$ |
| Thyroid | $0.0003 \pm 0.0003$ | $5.4 \pm 2.2$ | $0.271 \pm 0.110$ |
| Urinary bladder | $0.0833 \pm 0.0426$ | $51.4 \pm 30.3$ | $2.568 \pm 1.512$ |
| Uterus | $0.0015 \pm 0.0015$ | $9.4 \pm 6.9$ | $0.047 \pm 0.034$ |

*Total effective dose* **12.1 ± 2.2 µSv/MBq**

### Biodistribution and dosimetry

PET imaging started immediately upon tracer administration and consisted of a series of passes covering the whole body in seven bed positions. To capture the kinetics of the tracer and maximize image quality, the acquisition time per bed position was short at first (1 min per bed position) and increased over time (2, 4 and 8 min per bed position). Whole-body and brain PET images at four selected time points are shown in **Figure 2**. As it can be seen from the images and the accompanying time-activity curves (TACs), the tracer quickly distributed throughout the whole body including the brain. The maximum activity 8-14 min post-injection can be seen in the liver, kidneys, urinary bladder, spleen, stomach and brain. At 22-28 min post-administration, the highest activity was present in the kidneys, biliary duct and urinary bladder. After 60 min, most of the activity had cleared from the organs and accumulated in the urinary bladder. Fat was the tissue type with the lowest activity and no activity accumulation was observed in bone indicative of no defluorination. The biodistribution seen in the whole-body images indicates rapid clearance primarily through the kidneys with a small amount of radioactive tracer or metabolites excreted slowly through the hepatobiliary system.

**Fig. 2.**
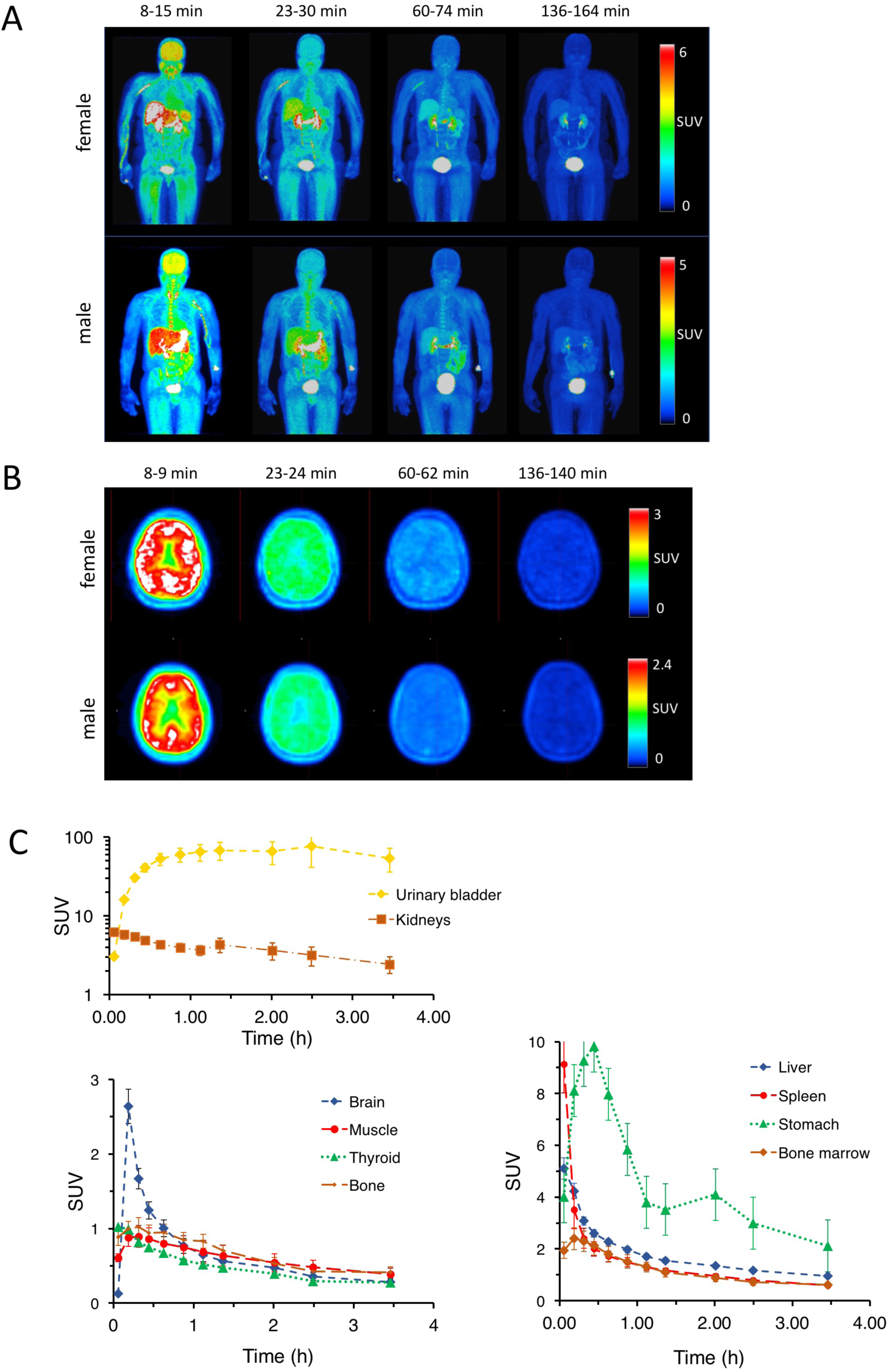
[^18^F]3F4AP in human subjects. **A**. Representative whole-body maximum intensity projections of a male and a female participant at different time points. **B**. Representative brain images of a male and a female participant. **C**. Averaged time-activity curves of selected organs.

Organ and whole-body dosimetry were calculated from the TACs using OLINDA software as described in the methods. Organs with the highest dose were the heart, urinary bladder and kidneys whereas the organs with greater contribution to the effective dose were the ovaries, urinary bladder and testes. The calculated average effective dose was 12.2 ± 2.2 µSv/MBq for the four participants and no differences were observed between male (11.2 ± 1.6 µSv/MBq) and female (12.9 ± 1.8 µSv/MBq) participants. The effective dose was significantly lower than the effective dose estimated from nonhuman primate studies (21.6 ± 0.6 µSv/MBq)[11] and the effective dose of other PET tracers ([^18^F]FDG ED = ∼20 µSv/MBq[14]). The lower ED is likely due to the faster clearance in humans than in nonhuman primates. **Figure 3A** shows averaged whole blood radioactivity concentration curves of the four human subjects and two nonhuman primates and indicates faster clearance in humans. Interestingly, the blood concentration measured from human blood samples is very similar to the concentration in blood obtained from a VOI placed in the left ventricle of the heart suggesting that the blood concentration obtained from PET images may provide a good input function for pharmacokinetic modeling (**Fig. 3B**).

**Figure 3.**
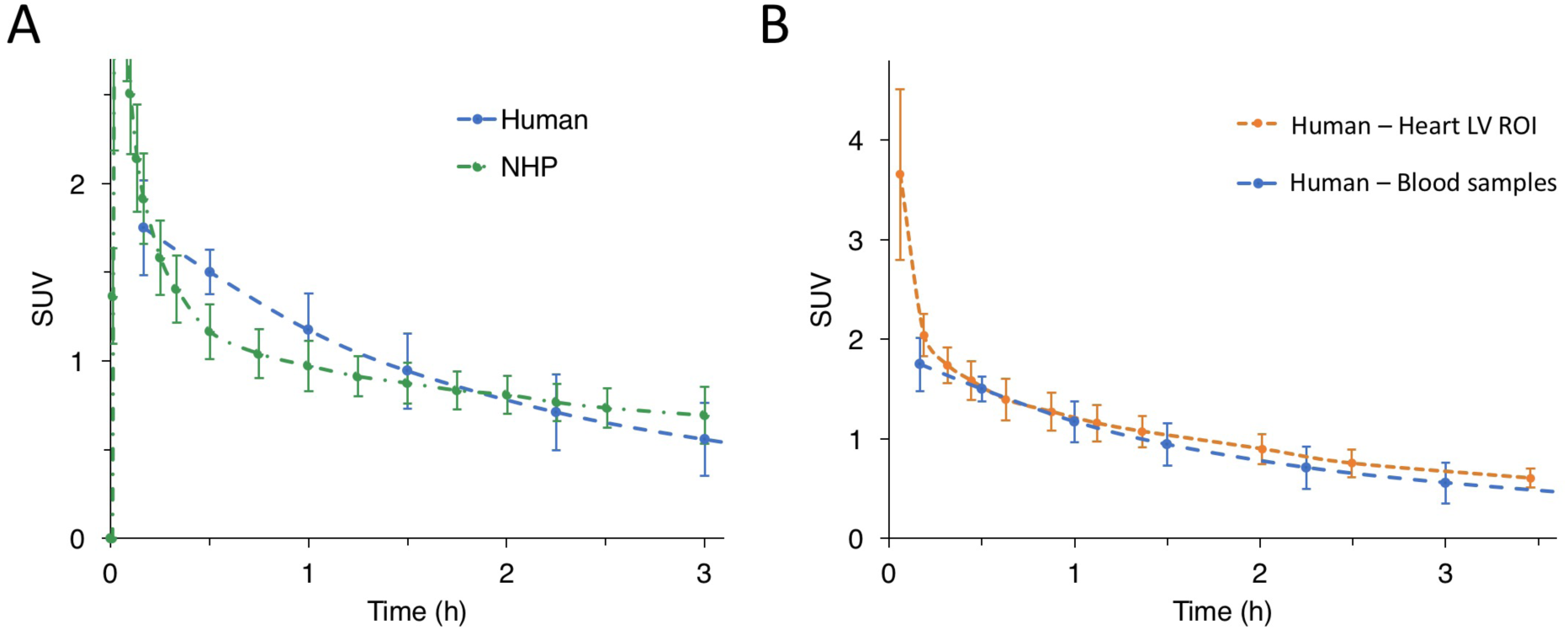
Clearance of [^18^F]3F4AP. **A**. Whole blood time-activity curves in human *vs*. nonhuman primate. **B**. Comparison of radioactivity concentration in human blood measured by gamma counting venous samples and the concentration derived from an ROI placed in the heart left ventricle in the PET images.

### Safety assessment

No significant changes in vital signs (temperature, blood pressure and oxygen saturation) were observed before and after the scan. In addition, no significant changes were noted in blood CMP results and electrocardiogram results obtained within 30 days before and 30 days after. The tracer and the imaging procedure were well tolerated by all the participants and no adverse events occurred during the scan.

## Discussion

We present results from the first study with [^18^F]3F4AP in human research subjects. The goal of this study was to assess the biodistribution, radiation dosimetry and safety of [^18^F]3F4AP in healthy volunteers. The study included four healthy adults (two males and two females). The biodistribution and calculated effective doses across subjects were found to be very consistent justifying scanning a small number of participants. The tracer was found to distribute widely throughout the body including into the brain and clear quickly primarily via renal excretion. The clearance of the tracer was faster than expected from a previous study in rhesus macaques as evidenced by a faster drop in blood concentration compared to NHPs. This faster clearance resulted in lower organ doses and a lower effective dose than predicted from NHP studies. The reasons for the faster clearance may be due to the effects of anesthesia since monkeys were imaged under anesthesia whereas humans are imaged awake and will be investigated in the future. Importantly, and as expected, the tracer and study procedures were well tolerated among study participants.

## Data Availability

The datasets generated during and analyzed during the current study are available from the corresponding author on reasonable request.

## Acknowledgments

The authors thank David Lee, Jr. for providing the fluorine-18 for the radiotracer synthesis. Prof. Brian Popko from Northwestern University is also acknowledged for supporting the study.

## Author contributions

PB, MQW, NJG, MDN and GEF designed the study. JN, DRV, KMRT, PAR and DLY established the radiosynthesis method, produced the tracer and performed quality control. JAK, MTMS recruited and scanned the participants. MTMS and NJG collected the blood samples and analyzed the blood data. RL supervised the safety aspects of the study. PB, DYL, JAK and GEF prepared the IRB and IND documentation. PB and MQW analyzed the data and wrote the manuscript, which was reviewed and approved by all the authors.

